# ApharSeq: An Extraction-free Early-Pooling Protocol for Massively Multiplexed SARS-CoV-2 Detection

**DOI:** 10.1101/2020.08.08.20170746

**Authors:** Alon Chappleboim, Daphna Joseph-Strauss, Ayelet Rahat, Israa Sharkia, Miriam Adam, Daniel Kitsberg, Gavriel Fialkoff, Matan Lotem, Omer Gershon, Anna-Kristina Schmidtner, Esther Oiknine-Djian, Agnes Klochendler, Ronen Sadeh, Yuval Dor, Dana Wolf, Naomi Habib, Nir Friedman

**Affiliations:** Silberman Institute of Life Science, Hebrew University of Jerusalem, Jerusalem 9190401, Israel; Rachel and Selim Benin School of Computer Science, Hebrew University of Jerusalem, Jerusalem 9190401, Israel; Edmond and Lily Safra Center for Brain Sciences, Hebrew University of Jerusalem, Jerusalem 9190401, Israel; Hadassah - Hebrew University Medical Centre, Jerusalem 9112001, Israel; The Lautenberg Centre for Immunology and Cancer Research, IMRIC, Faculty of Medicine, The Hebrew University of Jerusalem, Jerusalem 9112001, Israel; Department of Developmental Biology and Cancer Research, IMRIC, Faculty of Medicine, The Hebrew University of Jerusalem, Jerusalem 9112001, Israel

## Abstract

The global SARS-CoV-2 pandemic created a dire need for viral detection tests worldwide. Most current tests for SARS-CoV-2 are based on RNA extraction followed by quantitative reverse-transcription PCR assays. While automation and improved logistics increased the capacity of these tests, they cannot exceed the lower bound dictated by one extraction and one RT-PCR reaction per sample. Multiplexed next generation sequencing (NGS) assays provide a dramatic increase in throughput, and hold the promise of richer information including viral strains, host immune response, and multiple pathogens.

Here, we establish a significant improvement of existing RNA-seq detection protocols. Our workflow, ApharSeq, includes a fast and cheap RNA capture step, that is coupled to barcoding of individual samples, followed by sample-pooling prior to the reverse transcription, PCR and massively parallel sequencing. Thus, only one non-enzymatic step is performed before pooling hundreds of barcoded samples for subsequent steps and further analysis. We characterize the quantitative aspects of the assay by applying ApharSeq to more than 500 clinical samples in a robotic workflow. The assay results are linear, and the empirical limit of detection is found to be Ct 33 (roughly 1000 copies/ml). A single ApharSeq test currently costs under 1.2$, and we estimate costs can further go down 3-10 fold. Similarly, we estimate a labor reduction of 10-100 fold, automated liquid handling of 5-10 fold, and reagent requirement reduction of 20-1000 fold compared to existing testing methods.

## Introduction

A novel Coronavirus, SARS-CoV-2, has infected over 25 million people, as of late September 2020, with almost one million related deaths caused by COVID-19^1^. There is a consensus that testing is paramount for the containment of the pandemic^2^, yet widespread testing in most countries is lacking. At the time of writing this manuscript, multiple countries are undergoing a “second wave” of infections, highlighting the need for population-level screens for prolonged durations, effectively requiring a billions of tests world-wide.

The current benchmark for SARS-CoV-2 testing is a panel of RT-qPCR tests approved by various institutions typically applied to nasopharyngeal swab samples^3^. The swabs are mixed into a lysis buffer (either directly or first into transport buffer and subsequently mixed with a lysis buffer), and undergo RNA extraction and RT-qPCR. Generally, in these tests, samples with cycle threshold (Ct) lower than 35 are considered positive. While this test is sensitive and specific, qualified labor and the shortage of required equipment and reagents have proven to be limiting factors at different stages of the pandemic^4^. Specifically, the testing capacity is limited since each sample is treated as a separate qPCR reaction with a fixed reaction time.

In the last decade, next generation sequencing (NGS) has replaced RT-qPCR and microarrays as the assay of choice for quantifying RNA molecules. During the pandemic, different groups suggested NGS-based assays to measure the existence and abundance of the viral genome in samples^5,6,7^. In addition to detection and quantification, these assays can provide strain-specific sequence information as events are unfolding, providing epidemiologists with data to analyze the contagion propagation through the population^8,9^. Similarly, by assaying the RNA from host cells, aspects of the immune response in infected individuals can be unmasked^10^, providing potentially crucial information for patient treatment, research, and policymakers.

Here, we propose a general modification of current RNA-seq protocols that allows for pooling of barcoded samples prior to reverse transcription - Amplicon Pooling by Hybridization And RNA-Seq (ApharSeq). This improvement is especially pertinent for large scale testing as it reduces the need for labor and reagents by orders of magnitude. Briefly, we show (Figure 1) that we can introduce barcoded and target-specific reverse transcription primers to the samples, allowing them to hybridize to target RNA molecules already in the lysis buffer, or after a brief RNA cleanup step. After several minutes of hybridization with the barcoded primers, we capture the sample RNA onto beads, washing away excess primers and the chaotropic lysis buffer. Importantly, the primer-RNA hybrids are preserved during this step. The bead-bound RNA is isolated, and samples are pooled to a single tube to undergo reverse transcription from the primers that remain hybridized to their targets. Finally, the pool undergoes library PCR and sequencing. Viral molecular counts per sample are determined by the sample-specific barcodes and unique molecular identifiers introduced at the very beginning of the protocol. We demonstrate that cross-sample contamination in this workflow is negligible, and determine sensitivity to be ~800-1600 copies/ml, comparable to existing FDA and EU approved tests^11^.

**Figure 1 -.**
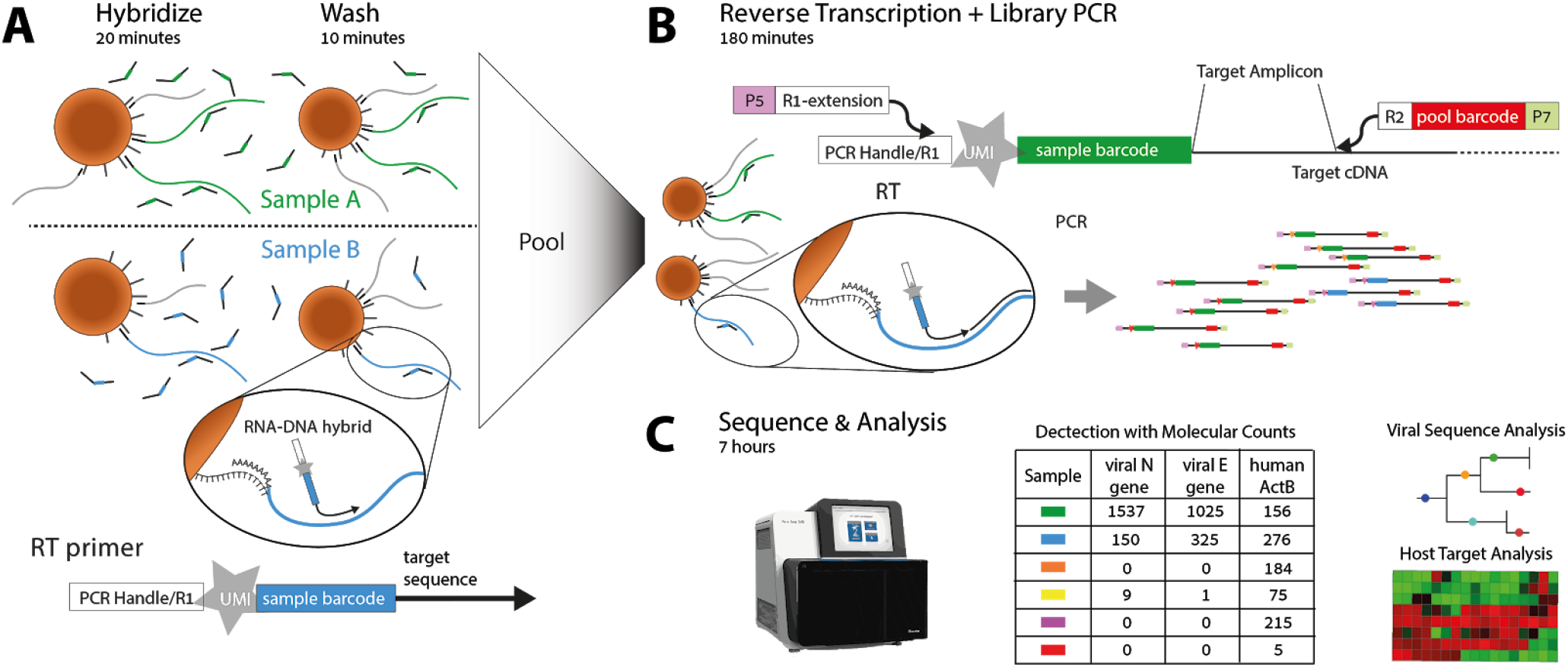
ApharSeq overview. **A)** Barcoded and uniquely-identifiable RT primers are hybridized to samples in transport/lysis buffer or after a quick buffer replacement step. Paramagnetic beads are used to capture RNA and wash excess primers. **B)** Beads are pooled and RNA undergoes an RT/PCR reaction with pre-hybridized target-specific primers to generate a sequencing library. **C)** Libraries are sequenced and analysed, PCR duplicates are collapsed to molecular counts for detection and potentially more elaborate analyses, e.g. contact tracing by viral sequence analysis, host physiological state by mRNA analysis.

## Results

### A simple and quick RNA capture step

Lysis buffers contain protein denaturation and degradation reagents, chaotropic guanidium thiocyanate, as well as detergents. Therefore, enzymatic reactions such as reverse transcription typically require RNA extraction from the lysis buffer in which the samples arrive. The Sars-CoV-2 genome is a poly-adenylated 30 kb RNA molecule, therefore we tested two home-made bead-based RNA cleanup protocols, SPRI and polyT capture^12^, that are cheap, simple, amenable to automation, and take less than 30 minutes for 96 samples. In terms of RNA yield, the performance of both approaches is within a ±50% range of a widely used commercial kit (see supplementary note on RNA capture; Figure 2A; Figure S1). Preliminary tests showed that both approaches can be used for ApharSeq (Figure S1D), but we focused on the polyT-based variant. Note that we do not elute the RNA from the polyT beads, rather we use the beads to capture the RNA and continue with the bead-bound material to the next step of the protocol.

**Figure 2:**
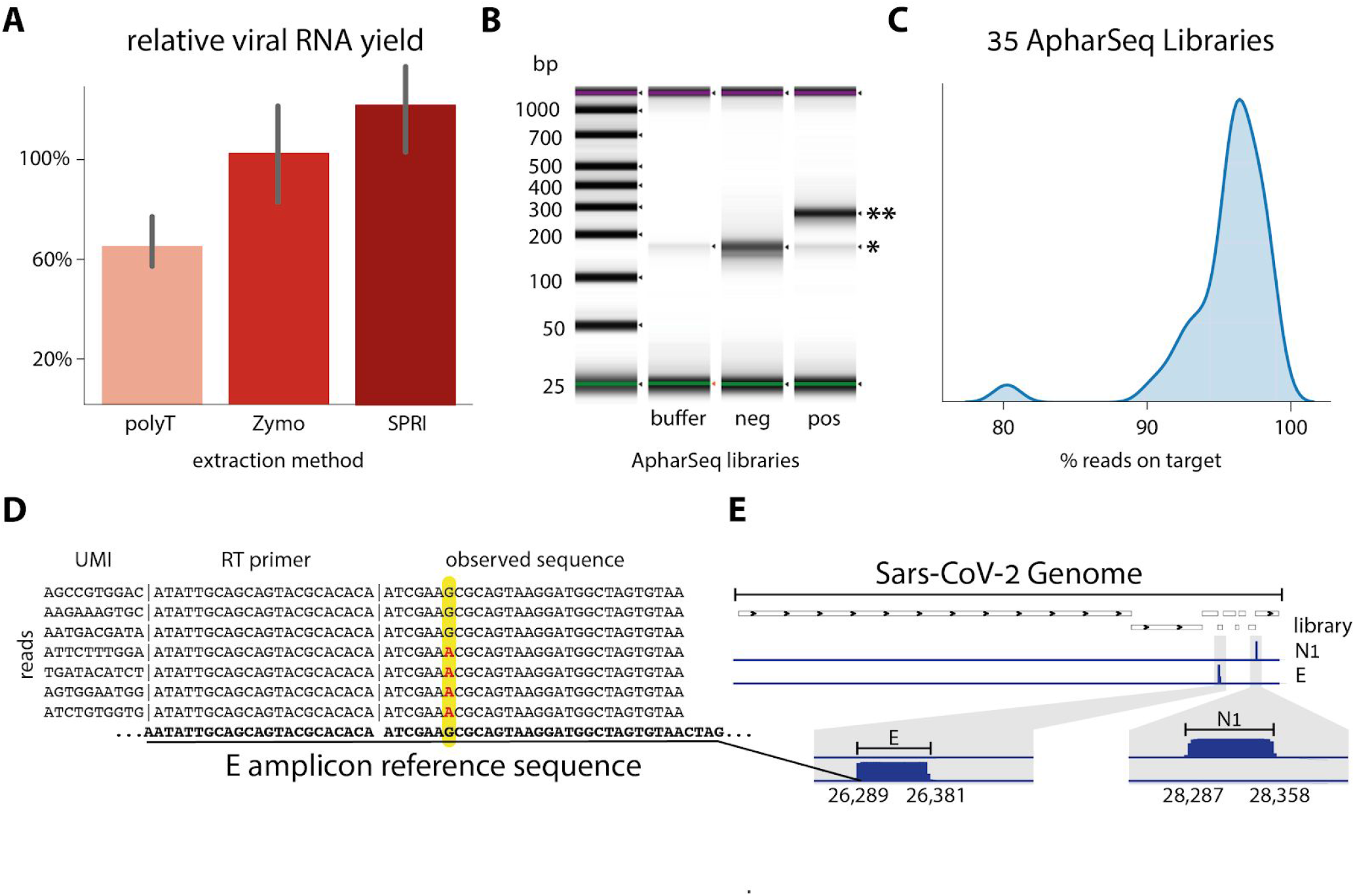
The ApharSeq pipeline generates specific sequencing libraries. **A) RNA capture** by homemade polyT and SPRI paramagnetic beads is efficient (~60-120% compared to Zymo kit), and quick (<30’ for a 96 plate on a robotic system). See supplementary note on RNA capture. **B) Sequencing libraries** of the viral envelope amplicon (E) are specific to positive samples (* primer dimer, ** expected amplicon: 269 bp). **C) Libraries are highly specific**: >75% of libraries have more than 95% of reads mapped to the expected sequence. **D) Observed sequences** conform to the reference genome in >99.9% of reads (not shown). However, in at least one sample we observe a sequence variation in reads from the E amplicon at 26,353 (G to A). Observed SNP is in more than 90% of reads of each UMI shown (~200 reads each), excluding the possibility of sequencing errors. **E) Genome browser view** of N1 and E amplicon libraries highlight target specificity

### Barcoded RT primers added to lysed samples prime RT reactions

We designed barcoded RT primers for the viral E gene (reverse), as it appears in the WHO panel^13^. The primer includes a 10bp barcode and a 10bp unique molecular identifier to allow for single-molecule counting^14^ (Figure 1A, methods). Each sample is hybridized to primers with a different barcode (Figure 1A), effectively identifying the RNA for the remainder of the protocol. The bead-bound RNA is then washed and reverse transcribed to generate sample-labeled cDNA.

To evaluate the efficacy of primer-RNA hybrid formation and stability through the cleanup stage, we designed a qPCR reaction targeting the generic PCR handle on the RT primer and the amplicon target sequence. This assay allows us to quantify the number of hybrids that survive the washes and succeed in generating a cDNA molecule. Using the qPCR assay, we established that RT primers indeed remain hybridized during the RNA capture and initiate reverse transcription reactions (Figure S2). Furthermore, we used this simple assay to run several optimizations for the first steps of the protocol and improve the yield significantly (Figure S2).

### Sequencing library preparation

The next step in the ApharSeq protocol is to generate sequencing libraries. This is achieved in a single PCR step, amplifying the RT primer from the generic sequencing handle and from the forward amplicon-specific primer, extending the amplicon with Illumina-compatible handles (Figure 1B). When we applied this PCR to positive and negative samples we consistently obtained amplicon-specific libraries only in SARS-CoV-2-positive samples (Figure 2B). When sequenced on a next generation sequencing platform, these libraries yield highly specific results with >95% of reads aligning to expected viral target sequence in positive samples (Figure 2C). Importantly, sequencing the insert provides direct evidence for the presence of the virus in samples, bolstering confidence in the assay’s results. Observing the target sequence directly allowed us to identify viral sequence variations in some cases (Figure 2D).

### Cross-Sample Contamination is minimal

When pooling samples early on in the protocol, the main concern is that RNA molecules will be erroneously tagged due to residual free primers, or due to other artifacts during RT, PCR, or sequencing. To test cross-contamination levels in the RT stage, we hybridized positive (Ct 26) and negative samples with two differently barcoded primers, pooled them, performed RT and tested the amount of cross-contamination by barcode-specific qPCR (Figure 3A and 3B). We find that the pooled negative sample is indistinguishable from the unpooled negative sample, suggesting cross contamination is negligible.

**Figure 3:**
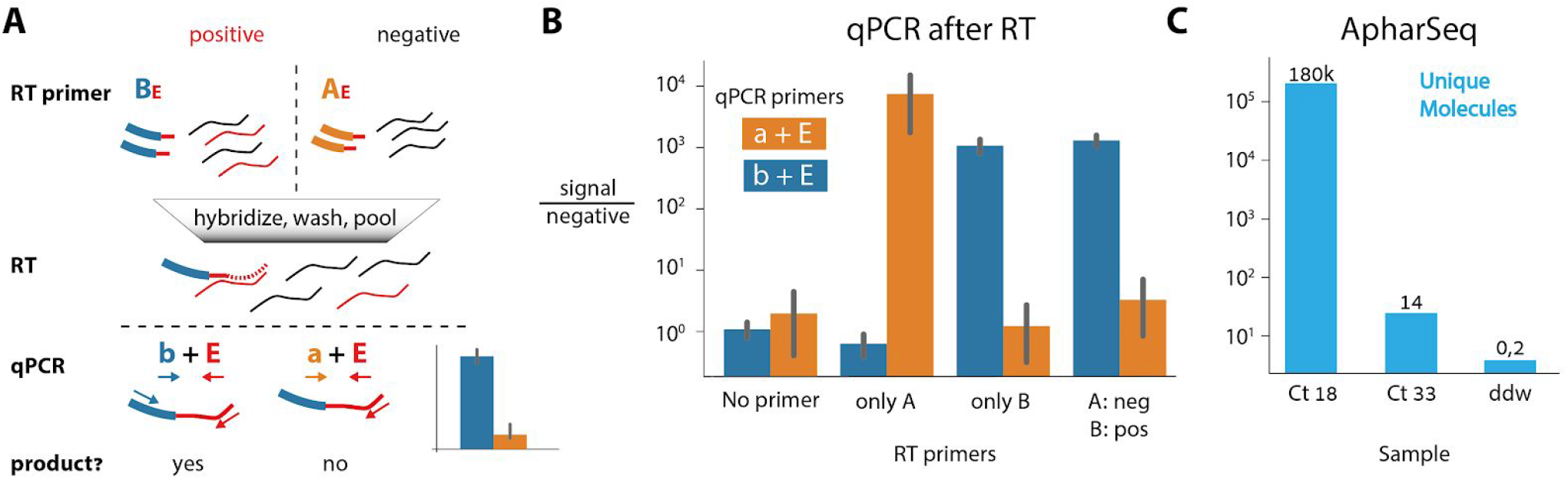
Minimal Cross-contamination in ApharSEQ. **A) qPCR cross-contamination assay -** Different RT primers (uppercase AE/BE) targeting the same locus (viral E amplicon) are hybridized to a positive/negative sample, then samples are pooled, reverse transcribed, and RT-primer-specific qPCR primers (lower case a/b) are used in qPCR to detect successful RT reactions on the viral target. **B) qPCR results** in fold change relative to no template control. **C) Minimal cross contamination test in ApharSeq libraries** as quantified by unique molecules detected in a pool of four samples with Ct 18, Ct 33 and two negative controls. Numbers above the bars are the counts of unique molecules in each sample.

Next, we examined potential cross-contamination during PCR or sequencing^15^. We subjected four samples - Ct18, Ct33, and two negative controls (ddw) to ApharSeq. We hybridized the barcoded primers, and then pooled the samples prior to RT and PCR (Figure 3C). Again, we find that barcodes that were hybridized to negative samples have at least 50,000-fold less reads than those that were hybridized to the high Ct positive sample. These results are not unique to the polyT-based capture, and were qualitatively replicated using SPRI-based RNA cleanup (Figure S1D). We conclude that cross-sample contamination is a minor issue in ApharSeq.

### ApharSeq is quantitative and sensitive

To evaluate the dynamic range of ApharSeq, we titrated a positive sample into lysis buffer and generated samples that span the Ct range ~23-31 in 320 μl. We applied ApharSeq to these samples in a pool and as individual samples (Figure 4A). We find that the number of unique molecules scales linearly with the input (p-value < 0.001; Figure 4B). Accounting for observed background in negative samples, we predict the limit of detection to be ~Ct 35.7. Importantly, the linear titration curve in the case of the pooled samples highlights the minimal interaction and contamination between pooled samples.

**Figure 4:**
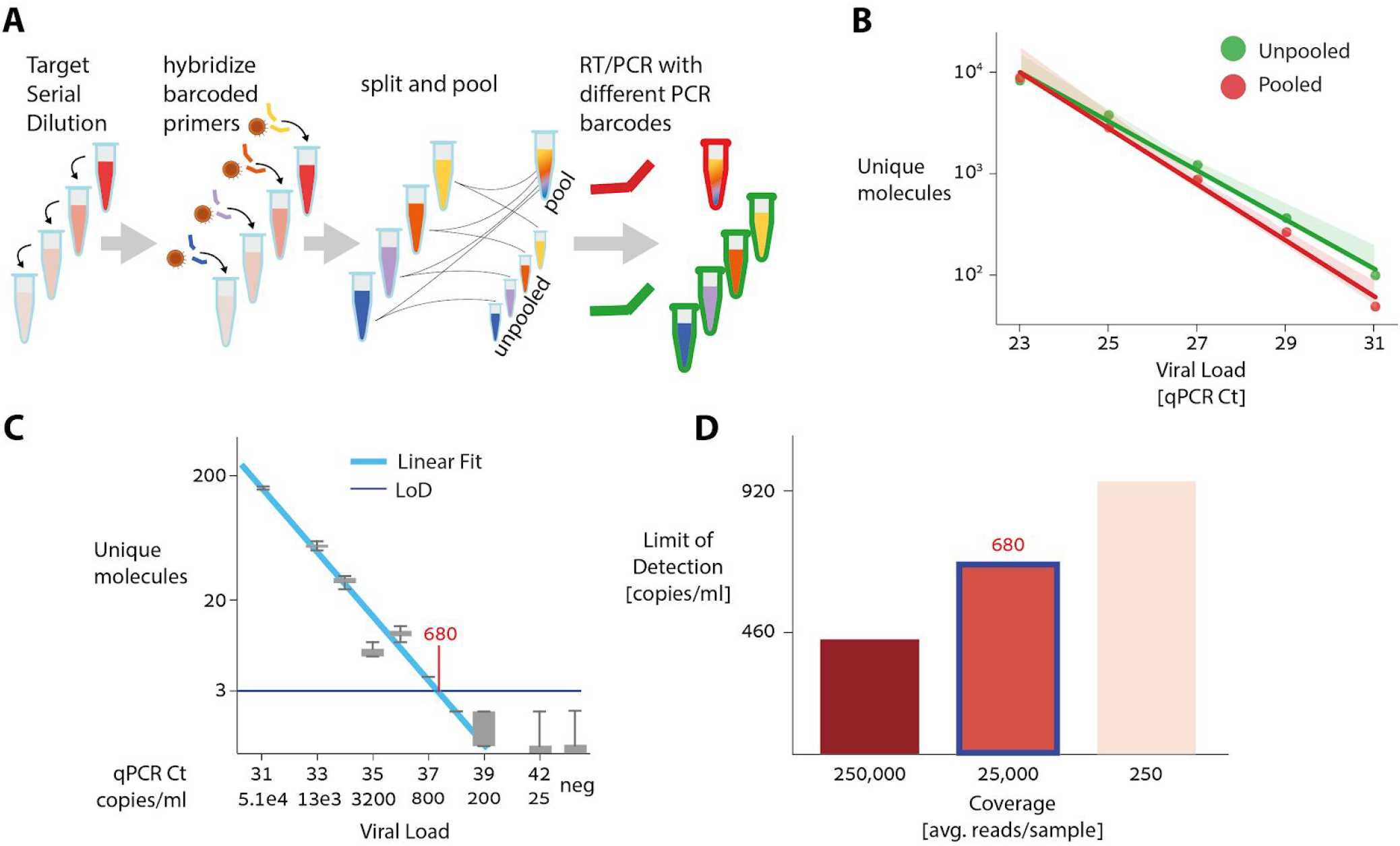
ApharSeq Sensitivity. **A) Target titration experimental scheme** SARS-CoV-2 positive sample is diluted in lysis buffer. Diluted samples are hybridized to barcoded ApharSeq primers (interior tube color), and split so they can be assayed separately or pooled. Samples are subjected to PCR with different barcodes (red/green) to distinguish their treatments. **B) Assay is linear** in both cases (p value < 0.001). The linearly extrapolated LoD for pooled samples is ~Ct 35.7 and is 4 times lower than the LoD for the single samples (methods). **C) Low target titration experiment**, in which we also introduced a viral target control for quantification (methods) shows the actual LoD is ~680 molecules/ml (Ct ~37.4). **D) Limit of detection** as a function of Sequencing Depth as derived by down-sampling the sequencing reads (methods). Highlighted bar (~25,000 reads per sample) corresponds to data in C.

We next tested the limit of detection directly, by performing another pooled titration experiment with highly diluted samples with Ct range 30-42 (Figure 4C), while adding a quantified reference (methods). Using these quantified controls, we estimate the end-to-end capture rate of ApharSeq at ~1.5% (observed 33 and 14 molecules out of an input of ~2000 and ~1000 molecules, respectively). Similarly, we could also calibrate the titration curve from Ct units to molecular counts, and found the limit of detection to be 450-900 molecules/ml, depending on sequencing depth (Figure 4D), threshold selection, and input volume used (methods).

### Multiple Target Assay

A major advantage of sequencing-based assays is the capacity to capture and readout a large number of targets from the same sample^16^. As a first step towards a multi-target assay, we aimed at multiplexing two targets. We started by designing RT and PCR primers for the viral N1 amplicon, as it appears in the CDC panel^17^, and used it in conjunction with the E amplicon RT and PCR primers. We applied ApharSeq to a positive sample with each primer separately or with both primers together (Figure 5A). The results of individual and multiplexed amplicons are almost identical (Figure 5B), suggesting that the viral targets are largely independent and can be probed simultaneously to improve confidence, and potentially improve sensitivity. Notably, the N1 amplicon yields roughly 2-3 fold more molecules, consistent with previous reports^18^.

**Figure 5:**
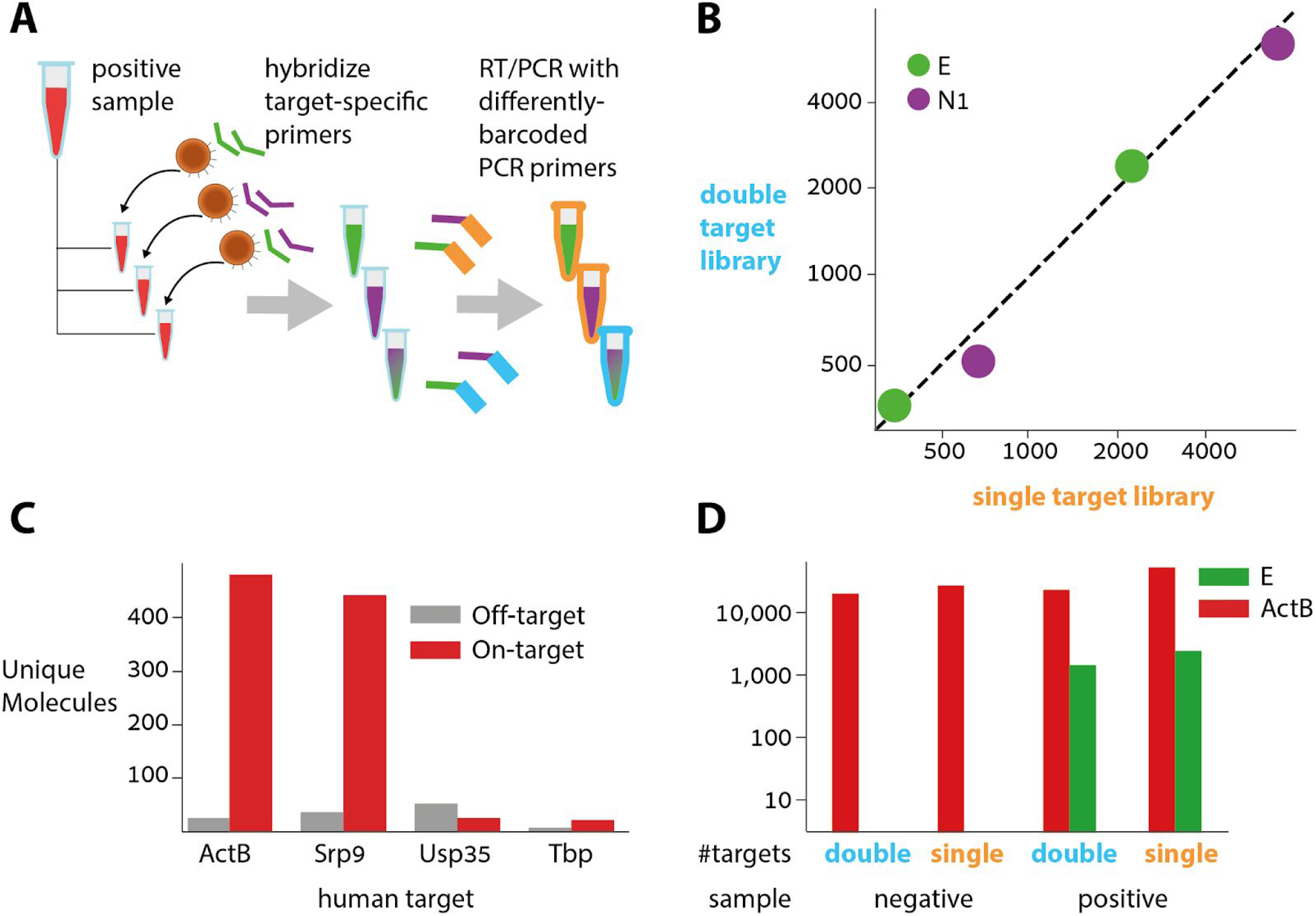
Multi-target ApharSeq libraries. **A) Multiple target assay scheme**. A positive sample was split and hybridized with E, N1, or E+N1 RT primers. These samples underwent ApharSeq with differently-barcoded PCR primers to delineate reads from the double/single hybridization conditions. **B) Target Multiplexing Results**. ApharSeq libraries for high/medium viral load samples for the N1 and E amplicon separately (x-axis) yield similar counts to a single ApharSeq library for both targets (y-axis). Units are unique molecular counts. **C) Human Target Tests** for multiple genes on a pool of negative samples highlights potential specific targets at different expression levels (see all targets tested in Figure S3). **D) Human and viral targets multiplexed** in the same ApharSeq library (“double”) allow for internal control in negative/positive samples. Units are unique molecular counts.

Next, as an internal control, we designed primers for several human transcripts with varying expression levels^19^ and after a preliminary test (Figure 5C, Figure S3) we decided to continue with a β-Actin amplicon as it is also used in an approved detection kit^20^. Subjecting positive and negative samples to the ApharSeq pipeline with primers targeting viral E and human ActB amplicons produced sequencing libraries, albeit with slightly reduced yields (Figure 5D). Importantly, qPCR tests on mixed libraries showed that titration of the human-specific primer in the PCR affects the human/viral amplicon ratio accordingly, allowing for calibration of the number of reads allocated to each target in a multi-target library (Figure S3).

### Robotic ApharSeq on clinical samples

Finally, we wanted to validate that ApharSeq can scale to hundreds of samples. To this end we developed and tested a robotic protocol on a Tecan liquid handling station (Methods). With our current preliminary protocol, a single 96 sample plate is processed in 40 minutes, and then can be pooled to a single tube for RT-PCR.

To test ApharSeq in a real setting we obtained dozens of positive samples and hundreds of negative samples. We randomly assigned these samples to six 96 sample plates, and added positive and negative standards to each plate (Figure 6A, Methods). These plates were split in two - one half underwent a standard RNA extraction and quantification with a RT-qPCR kit, and the half underwent the ApharSeq protocol (Figure 6A, Methods).

**Figure 6:**
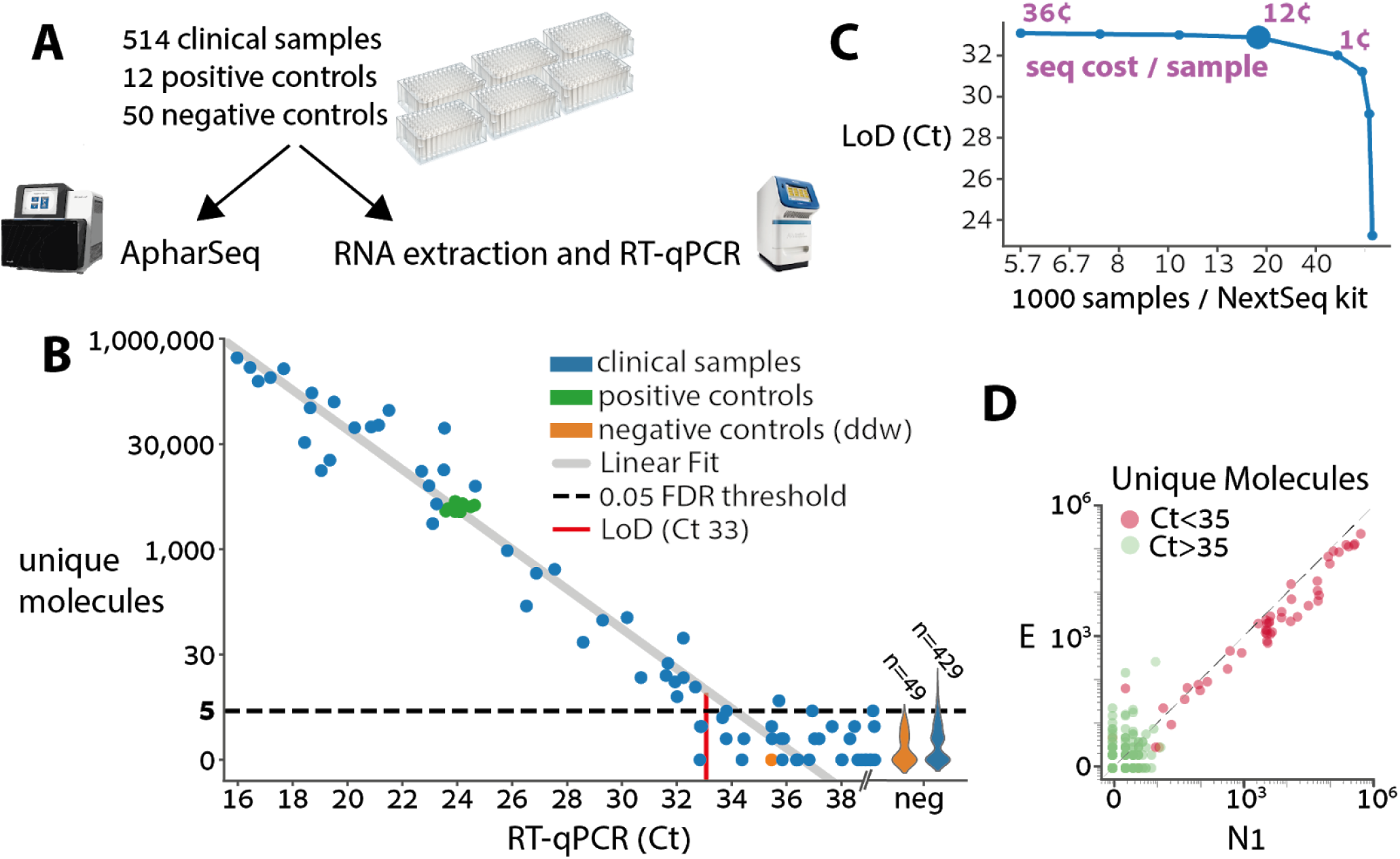
Robotic test of hundreds of clinical samples. **A) Experimental design** for clinical samples. Samples were randomly assigned to 96 sample plates with ddw negative controls in each plate and two standard positive controls derived from a clinical sample that was diluted in negative samples as to suffice for multiple tests. The plates were then split and subjected in parallel to ApharSeq or to the standard clinical pipeline at Hadassah hospital (Zymo RNA extraction and BGI RT-qPCR). **B) ApharSeq is highly quantitative** as depicted when the number of unique N1 molecules observed in each sample (y-axis) is plotted against the measured Ct for the same sample. Limit of detection (LoD) was determined by controlling the false positive rate to be 5%, and assuming Poisson noise (methods). Violins on the right depict the distribution of unique molecules observed for the samples that were not detected in the RT-qPCR. Note the high reproducibility among the positive control samples (low y-axis variance). **C) Downsampling analysis** of the sequencing data demonstrates that samples are sufficiently sequenced, and that there is a minimal decrease in sensitivity (y-axis) when sequencing depth decreases down to ~25,000 reads per sample which is equivalent to ~16,000 samples in a single NextSeq run (larger blue marker). Purple numbers indicate the sequencing cost per sample in selected sequencing depths. **D) Viral amplicon correlation within samples**. Unique molecules of the E and N1 amplicons are plotted per sample, demonstrating reproducibility. Colored by the Ct threshold currently used in the clinic.

The positive controls exhibit excellent reproducibility between different plates highlighting the stability and robustness of the protocol (Figure 6A). In addition, there was a clear quantitative agreement between the human internal control in the RT-qPCR assay and our ActB amplicon reads, with a similar amount of missing values in both assays (i.e. samples that will require re-testing, Figure S7). When we compare the Ct value of each sample to the amount of unique molecules we observe from the N1 amplicon there is a strong linear agreement (R2 0.95, p-value < 10-38) establishing that the assay is highly quantitative (Figure 6B). Similarly, we observe excellent correlation between the N1 and E amplicons, bolstering confidence in the results. In concordance with other reports^21^, we observe more unique molecules of the N1 amplicon in virtually all the samples (Figure 6D). A preliminary analysis of the results, in which we fit a zero-inflated poisson model to the negative samples, and a linear model to the positive samples, determined the LoD in our experiment to be Ct 33, which is equivalent to ~1000 copies/ml (Figure 6B, Methods). A sub-sampling analysis of the data shows that sensitivity is maintained down to a sequencing depth of ~25,000 reads per sample (Figure 6C). Importantly, a single person was able to process these ~600 samples in less than 4.5 hours with a single robotic system, and we believe this can be significantly improved.

Overall, we conclude that the robotic ApharSeq protocol works efficiently, with minimal cross-sample contamination, and is highly quantitative.

## Prospects for massive testing

The results presented in this manuscript establish the ApharSeq pipeline as a SARS-CoV-2 detection test. We believe that ApharSeq is uniquely poised to allow tests at a massive scale, for both the low cost and minimal labor it requires.

The cost for a single test is currently between 1$-4$ (when processing 16,000-1,000 samples, see supplementary note on costs), and the bulk of the costs are due to consumable plastics (tips, plates, etc, >60%), primer synthesis and beads (15%), and sequencing (10%). These prices can probably be reduced by a factor of 5-10 when the process is streamlined, and reagents are manufactured and purchased in bulk.

In terms of labor, the bottleneck is the first step that involves per-test hybridization and washes. While this can be performed manually in roughly 45’ for 96 samples with a multichannel pipette, the prefered setup includes an automated liquid handling station, in which case this step requires 20-40 minutes, depending on the sample volume tested and the specifics of the robotic setup.

The sequencing requirement for the test is reading a single pool barcode (8 bp) and at least 50 bases from read1 or 20 bp from read1 and 30 bp from read2. When we down-sampled the data in the titration experiment, we found that a change of x1000 in sequencing depth only incurs a 2-fold reduction in sensitivity. Additionally, using the Ct distribution observed in the clinic, we performed a simulation to estimate the false positive and false negative rate as a function of sequencing depth (Figure S5). In these simulations, when sequencing depth ranged from 10,000 to 100,000 reads per sample the false negative rate ranged from 4.5% to 0.2% respectively. We conclude that 50,000 reads per sample on average should suffice (Figure 4D, figure S5). In a similar vein, a downsampling analysis of the clinical test data showed that 25,000 reads per sample are sufficient. This means that a single NextSeq 500/550 run with 400×10^6^ reads suffices for processing 8,000-16,000 samples, and a NovaSeq S2 100bp run with 8×109 reads allows the processing of 160,000-320,000 samples.

While highly informative and rich, the main drawbacks of sequencing-based assays are twofold - the requirement for specialized and expensive equipment, and the slow generation of data (e.g. a NextSeq 55bp run requires ~6.5 hours). While a 12-18 hour turnaround time is probably prohibitive for emergency testing, we believe that for large scale and routine population screens it is reasonable. Further, there are potential optimizations and workarounds that can reduce the sequencing runtime by a factor of 2-3^22^.

Importantly, we have preliminary results (not shown) that pooled RNA can be maintained for at least 24 hours in a standard RNA preserving buffer, and therefore samples can be processed in different facilities and collected to a central sequencing site for the last steps of the protocol (Figure 7).

**Figure 7:**
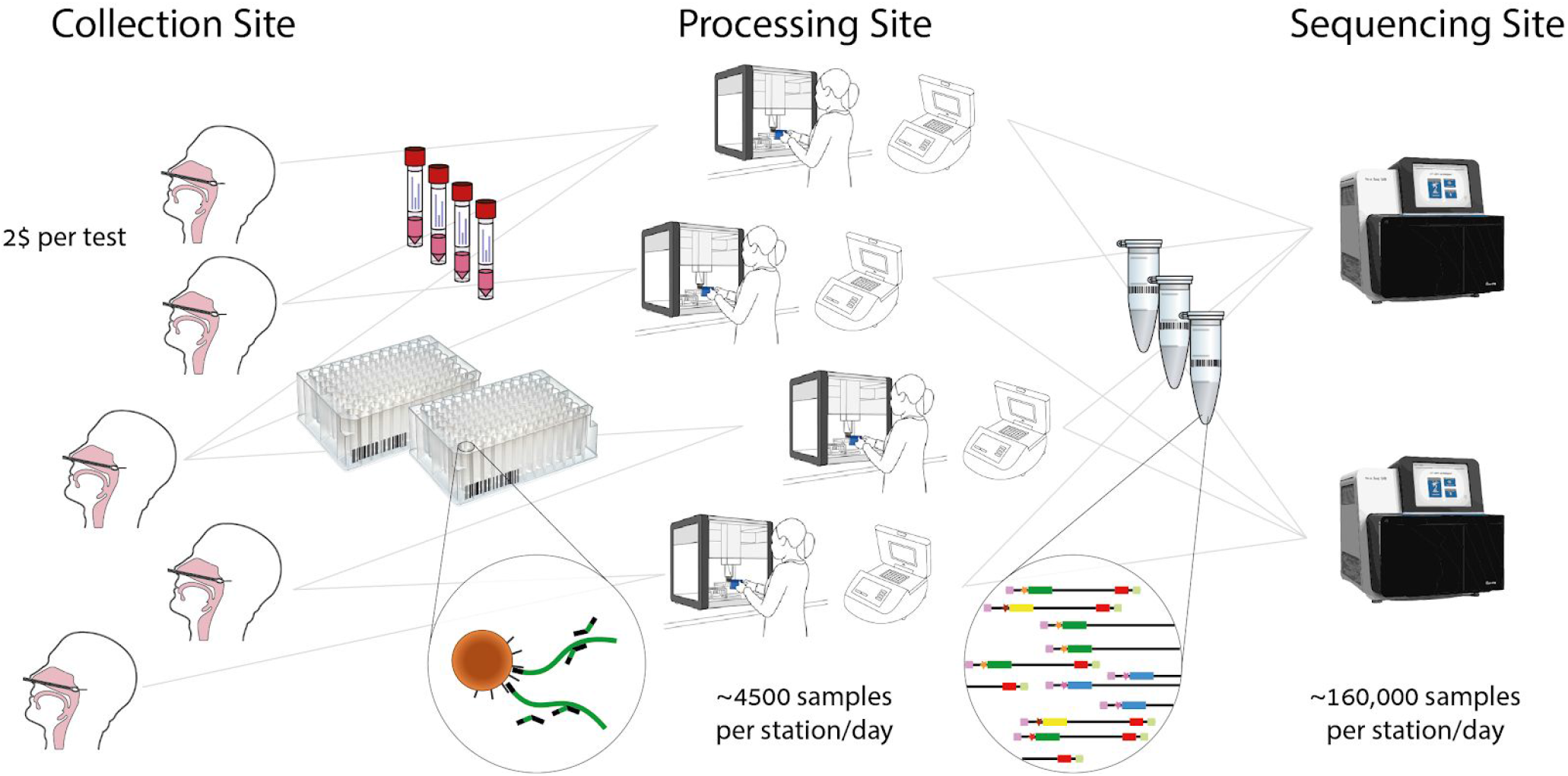
High Throughput testing at less than 2$ per test. We propose a division of labor scheme where any facility that has an automated liquid handler and a PCR block can function as a first step in the processing pipeline. A single 96 plate is processed and pooled every ~30 minutes in every such station. These pooled batches can be collected to a central sequencing facility that pools them further for cost-effective sequencing. If collection into plates is not feasible, there are dedicated automated liquid handler stations for re-organizing tubes into plates, as is already the case in most testing facilities.

Since the main equipment required for the application of ApharSeq already exists in many diagnostic facilities, we envision a gradual transition from the qPCR technology to sequencing-based assays by partial re-purposing of labs. For example, a facility with five liquid handlers can process upto ~2000 samples in two hours^12^, and then get back to its usual testing qPCR program. The pools generated during these two hours from four such facilities can be collected to a central sequencing center.

Once such advancements are made and ApharSeq proves useful and informative it can be used as an alternative avenue in case of supply shortages, or diversify the types of tests a single facility can provide for different use cases, namely high-throughput non-urgent population screens vs. lower-throughput urgent testing.

## Discussion

Here we propose ApharSeq, an early-pooling protocol for the detection of the SARS-CoV-2 virus in clinical samples using next generation sequencing. While sensitive RT-qPCR assays are the backbone of testing in the current pandemic, they are lacking sequence information that may be crucial to trace infection chains, and more importantly, they are difficult to scale up.

The current second wave of infections highlights that global testing and tracing efforts require an orders-of-magnitude scale-up. We believe that our approach, and future improvements and modifications thereof, might form the basis for such an improvement of several orders of magnitude relative to current testing strategies. Specifically, the ability to pool hundreds or thousands of samples as early as possible, without losing the sample-specific information, is a crucial improvement on all currently published NGS-based tests^6,23,24,25,26,27^. The early pooling approach proposed here, which might be applicable to other protocols, elicits proportional reductions in costs and labor that directly translate to higher throughput in testing.

While we established key properties of our approach, namely linearity, high-sensitivity, low cross-reactivity, and the potential for multi-target testing, there is still much to be done. Specifically, more optimization can improve efficiencies, shorten durations, reduce background noise and increase the sensitivity. Similarly, it is straightforward to introduce multiple internal synthetic controls that will provide clinicians with reproducible and informative assay measures and reliable results^7^.

Once the detection hurdle is surpassed and the relevant infrastructure and logistics are set into place, ApharSeq should be easy to extend to other more advanced applications in the context of the disease. Testing for co-morbid or confounding pathogens like flu viruses, amplifying viral variable regions to identify infection chains^28,29^, and monitoring the host immune response with key transcripts^30^ are just some of the potential applications of our generic approach. While these will incur some additional costs, we believe that for many applications the benefits will significantly outweigh the minor costs.

Finally, we note that the approach developed and validated here - hybridization of barcoded primers followed by early pooling - is of a general nature and can potentially be used to enhance existing protocols, including single cell and bulk RNA-seq protocols.

## Data Availability

All data and code will be made available upon request.

## Acknowledgements

We thank Michal Rabani for critical comments and support. We thank Michal Bronstein, Abed Nasereddin, Idit Shiff, Adi Turjeman, Netta Barak, and Moran Yassour for their help. This work was supported in part by the Rothschild Foundation. AC is an Azrieli scholar and would like to thank the Azrieli Foundation for their support.

## Methods

### RNA extraction benchmarking

Viral RNA was extracted from an in-vitro grown virus (Ct ~14) and serially diluted 1:25 in a negative sample. Each dilution was subjected to RNA extraction in three methods:

1. 400 μl sample with Quick-RNA MagBead (Zymo research), according to manufacturer’s instructions
2. 400 μl sample following the polyT capture described below
3. SPRI-based capture as published^12^ with modified volumes: 152 μl sample, 51 μl beads, 153μl PEG buffer, 122μl binding buffer

For all extractions, the titration curves were linear (not shown). Samples in the linear range were corrected for their dilution and collated to estimate the mean relative yield and error for each extraction method (Figure 2A).

### Primer Design

All oligos used in this study are provided in a supplementary excel spreadsheet.

### RT primers

RT primers consist of four main parts - from 5’ to 3’ - a general illumina handle (nextera R1), a 10 bp unique molecular identifier, a 10bp barcode, and a target-specific primer. After the first iteration of sequencing experiments, we decided to: a) interleave the UMI (U) and barcode (B), to avoid long stretches of the same nucleotide in the UMI sequence, and b) add to each primer a variable sequence of 0-2 N’s prior to the amplicon primer, to increase sequence complexity in each machine cycle:

- Nextera R1 sequence handle - **GCGTCAGATGTGTATAAGAGACAG**
- UMI/Barcode - **UUUUBBBBBUUUUBBBBBUU**
- Offset - **N{0,2}**
- Target primer -
- N1 **tctggttactgccagttgaatctg**
- E **atattgcagcagtacgcacaca**
- ActB **acagcctggatagcaacg**

Example of a specific primer (c410 - ActB RT primer):

**GCGTCAGATGTGTATAAGAGACAGNNNNACCTCNNNNTATCANNNacagcctggatagcaacg**

#### PCR primers

There are two different PCR strategies we employed during the development process - one-step and two-step PCR. The two step PCR is composed of a first step that amplifies the target molecules with an extendable handle, and in the second step barcode and the remaining Illumina sequences are introduced. The one-step reaction performs everything in a single reaction with a single long primer (~90 bp). A one-step reaction is more convenient, and is less prone to contaminations (see supplementary note on contaminations), however, it’s less modular. Specifically - the long primer contains a target specific sequence and a barcode, which means that a barcoded primer collection must be synthesized per target. The two step PCR decouples this dependency, which means that a single collection of barcoded primers can be used on any target, assuming a simple target-specific primer is used in the first step. Both approaches yielded similar results, and we are currently using the single-step reaction to avoid contaminations.

#### One-step PCR reaction

The PCR amplifies the generic handle on the RT primer on one side, and a target-specific sequence on the other side. Additionally, the PCR extends the amplicons to a sequencing library by adding the relevant flanking Illumina sequences. Including an 8bp barcode that marks the pool of samples amplified in the PCR reaction:

> Reverse primer (generic):
>
> CAAGCAGAAGACGGCATACGAGATGTGACTGGAGTTCAGACGTGTGCTCTTCCGATCT
>
> Forward primer (target-specific, and **indexed**)
>
> - N1 gaccccaaaatcagcgaaa
>
> - E acaggtacgttaatagttaatagcgt
>
> - ActB caccaactgggacgacat
>
> An example (c418 - ActB PCR primer):
>
> CAAGCAGAAGACGGCATACGAGAT**TCGGACTA**GTGACTGGAGTTCAGACGTGTGCTCTTCCGATCTcaccaac tgggacgacat

#### Two-step PCR reaction

The first step adds a target-specific handle on the forward side, and extends the generic handle in the reverse (RT) side of the amplicon. We do this with the Tn5-Rd1/Rd2 published (Illumina FC-121-1030) sequences:

> Reverse primer (generic):
>
> TCGTCGGCAGCGTCAGATGTGTATAAGAGACAG
>
> Forward primer (B6 - N1 PCR primer):
>
> GTCTCGTGGGCTCGGAGATGTGTATAAGAGACAGgaccccaaaatcagcgaaa
>
> The second PCR step extends the handles to a complete library with the Ad1.x and Ad2.x indexed primers as published^31^.

### ApharSeq Protocol

The detailed and complete protocol was published separately^32^. For convenience we briefly describe the protocol below. Since the protocol stabilized with time, some experiments are slightly modified relative to the current protocol. The supplementary material contains a list of experimental modifications per experiment shown indexed by figure panel.

#### Bead Preparation

We tested commercially available polyT beads (ThermoFisher dynabeads cat# 61002), or conjugated carboxylate coated beads (GE healthcare Sera-Mag SpeedBeads cat# 65152105050250), and followed the manufacturer conjugation protocol.

#### Hybridization and RNA purification

##### Option 1: Purification and hybridization on PolyT beads

This RNA purification protocol is based on a protocol for rapid isolation of mRNA^33^ with some modifications. Briefly, polyT conjugated beads were washed once and resuspended in binding buffer. The resuspended beads were mixed 1: 1 with the sample. After a hybridization period of 10 minutes at room temperature with periodic mixing, the supernatant is removed and the beads are resuspended in a 50 μl 1:1 mix of binding buffer and 10 pM barcoded RT primers. To denature RNA secondary structures, the samples were incubated at 72C for 2 minutes and immediately transferred to ice for at least 2 minutes. Samples were then incubated at room temperature for 10 minutes with periodic mixing to allow hybridization of RNA to the beads and to RT primers. Beads were resuspended in 450 μl wash buffer A and magnetized. Majority (380 μl) of the supernatant was removed and beads were resuspended in the remaining 70 μl of buffer A, and pooled. After pooling samples are washed once in buffer A, twice in buffer B, and can then be kept in RNA later until they are processed further. Preliminary tests show that RNA can be stored on the beads, in RNA later at 4C, for at least a week.

##### Option 2: Purification and hybridization on SPRI beads

RNA extraction with SPRI beads, followed our published protocol for RNA extraction^12^ with several modifications. Samples in lysis/transfer buffer were mixed with barcoded RT primers, then incubated at 72C for 2 minutes and immediately transferred to ice for at least 2 minutes. Samples were then mixed 1:1 with binding buffer (as above) and incubated at room temperature for 10 minutes with periodic mixing to allow primer hybridization. Next, samples were mixed 1:0.8 with home-made SPRI beads in PEG buffer^12^. Beads were washed twice with freshly made Ethanol 80%, air dried, and eluted in

DDW. This was followed by a second 0.8x SPRI cleanup to ensure the removal of any excess primers. At this stage samples were pooled to a PCR tube to undergo RT and PCR.

#### cDNA synthesis and Library preparation

25% of pooled beads are subjected to proteinase K treatment (Lucigen), washed, and undergo RT reaction with SmartScribe enzyme (SMARTScribe Reverse Transcriptase, Takara Bio) at 42C for one hour followed by incubation at 70C for 15 minutes. To elute the cDNA from the beads, the samples were incubated at 98C for 2 minutes, magnetized and the supernatant was transferred to a new tube and cleaned by SPRI beads x2 (Agencourt AMPure XP, Beckman Coulter). Illumina adaptors were added by PCR (KAPA HiFi HotStart ReadyMix, Kapa Biosystems, 30 cycles), and the DNA was purified using 1x SPRI.

### NGS data analysis

Reads were demultiplexed using bcl2fastq (version 2.20.0) and further processed by ad-hoc python scripts that are available as jupyter notebooks. We used the python API of UMI-tools to cluster UMI sequences. Specifically the “directional” option with the edit distance threshold set to three^34^

### Quantifying target molecules

To generate a quantitative polyA viral reference, we extracted RNA from a clinical sample and estimated the amount of molecules in this RNA extract to be 6000 molecules/μl using a synthetic viral sequence (Twist Bioscience SARS-CoV-2 RNA, MN908947.3) as a reference in a standard RT-PCR kit. We loaded two samples with 10 and 5 μl of this reference RNA in a total of 320 μl lysis buffer, and applied the ApharSeq protocol. Only 1/30 of the material underwent library preparation, which means that at most, we expect to see 2000, or 1000 molecules in the 10, 5 μl samples respectively. After UMI clustering we observe 33 and 14 molecules respectively, suggesting that we capture ~1.5% of molecules. In the same experiment, a sample corresponding to cycle 29.3 had a similar UMI count (32 molecules), allowing us to roughly calibrate the Ct units to target molecules / ml at Ct 29.3 = 6150 x 10 / 0.32 = ±190,000 molecules/ml. This number is much higher than the numbers reported in other publications (for example Ct 33.5 = 200 i.e. Ct 29.3 < 3500). If true, this means the LoD of our assay is actually *lower* than reported in this manuscript.

### Limit of detection determination

#### Titration LoD

For the high load titration (Figure 4B), a linear fit (python scipy.stats.linregress) was performed:

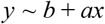

Where y is the log_10_(#UMIs) and x is the calculated Ct of the sample. Given this linear fit, we can extrapolate to the UMI detection threshold, which in this case was set to 3 (a conservative estimate). The fit statistics are:

| statistic | singles | pooled |
| --- | --- | --- |
| Slope (a) | -0.242 | -0.276 |
| Intercept (b) | 9.55 | 10.3 |
| p-value | 2.86e-4 | 2.53e-4 |
| R<sup>2</sup> | 0.99 | 0.99 |
| Ct @ $\log_{10}(3)$ | 37.48 | 35.7 |

For the low load titration (Figure 4C and 4D), we perform re-sampling of the data (x500 times):

> For factor in (1, 3, 10, 30, 100, 300, 1000):
>
> For each sample
>
> For each UMI in sample
>
> #sampled-reads(UMI) ← Poisson(#reads(UMI) / factor)

We then count the number of UMIs per (sample, factor, replicate) as the number of UMIs with #sampled-reads(UMI) > 0. Given these counts, we set the detection threshold as the minimal number of UMIs that is above 99% of replicates in the negative samples. Therefore this number varies with sequencing depth and the UMI background in the negative samples. We fit each sampled replicate of the data with a Poisson-noised exponent:

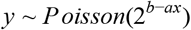

Where y is the number of observed UMIs and x is the calculated Ct of the sample. We then set the LoD per factor to be the maximal Ct such that 95% of replicates is above the LoD.

| resampling factor | Avg. #reads per sample | UMI threshold | LoD [Ct] |
| --- | --- | --- | --- |
| x1 | 247.7k | 3 | 38.0 |
| x3 | 82.6k | 3 | 37.8 |
| x10 | 24.7k | 3 | 37.4 |
| x30 | 8.26k | 3 | 37.2 |
| x100 | 2.47k | 3 | 36.9 |
| x300 | 826 | 2 | 37.2 |
| x1000 | 248 | 2 | 36.8 |

### Clinical Test LoD

Ct 35 was used as a cutoff value for positive/negative samples, as is currently used in the clinic.

We first determine the FDR threshold by fitting a zero-inflated Poisson mixture model (2 components + zero component) to the molecule counts observed in negative samples. Using this model we determine the theoretic threshold of detection, for a given false discovery rate. In this case, when FDR is set to 5% the threshold is 5 molecules.

We fit the positive samples with a linear model (slope = -1), assuming Poisson noise. Given the linear model, the FDR threshold (5 molecules), and assuming Poisson noise, the LoD is determined to be the maximal Ct in which the probability of obtaining a value lower than the threshold is lower than 5%.

We subsample reads from the data and repeat this analysis to every sequencing depth to obtain the LoD as a function of sequencing depth (Figure 6C).

### Clinical Tests

Clinical samples were collected in the clinical virology lab at Hadassah hospital. Positive samples were randomly assigned to plates and plate positions, as to have 7.5% positive tests on average. As a negative control we used ddw, and randomly assigned at least 6 such controls in each plate. As a positive control we diluted a positive sample in a pool of negative samples so that we had sufficient volume to allocate two positive controls to each plate. The plate positions of the positive controls were the same in all plates.

Plates were subjected to an automated script on an Evoware 100 Tecan system, and the pools were stored in RNAlater for further processing following the ApharSeq protocol.

